# Uncovering genetic associations in the human diseasome using an endophenotype-augmented disease network

**DOI:** 10.1101/2023.05.11.23289852

**Authors:** Jakob Woerner, Vivek Sriram, Yonghyun Nam, Anurag Verma, Dokyoon Kim

## Abstract

Many diseases exhibit complex multimorbidities with one another. An intuitive way to model the connections between phenotypes is with a disease-disease network (DDN), where nodes represent diseases and edges represent associations, such as shared single-nucleotide polymorphisms (SNPs), between pairs of diseases. To gain further genetic understanding of molecular contributors to disease associations, we propose a novel version of the shared-SNP DDN (ssDDN), denoted as ssDDN+, which includes connections between diseases derived from genetic correlations with endophenotypes. We hypothesize that a ssDDN+ can provide complementary information to the disease connections in a ssDDN, yielding insight into the role of clinical laboratory measurements in disease interactions. Using PheWAS summary statistics from the UK Biobank, we constructed a ssDDN+ revealing hundreds of genetic correlations between disease phenotypes and quantitative traits. Our augmented network uncovers genetic associations across different disease categories, connects relevant cardiometabolic diseases, and highlights specific biomarkers that are associated with cross-phenotype associations. Out of the 31 clinical measurements under consideration, HDL-C connects the greatest number of diseases and is strongly associated with both type 2 diabetes and diabetic retinopathy. Triglycerides, another blood lipid with known genetics causes in non-mendelian diseases, also adds a substantial number of edges to the ssDDN. Our study can facilitate future network-based investigations of cross-phenotype associations involving pleiotropy and genetic heterogeneity, potentially uncovering sources of missing heritability in multimorbidities.

## INTRODUCTION

Complex interactions between a variety of diseases can be explained by the presence of overarching groups of co-occurring phenotypes. Shared susceptibility between such diseases can be derived from common genetic, biological, or environmental factors. Indeed, diseases with comparable characteristics can occur simultaneously or sequentially with similar pathogenesis in a subject^1^. However, the best way to identify the contribution of genetic components to the etiology of such multimorbidities remains an open question. Due to the highly connected nature of diseases at the molecular level, it is necessary to concurrently examine not only phenotypes, but also the many genetic factors that could influence their pathological dynamics^2^. The field of network medicine offers an intuitive way of investigating the interactions between phenotypes^3^. Both global and local connectivity across multiple phenotypes can be explored through graph-based modeling and network representation. In particular, the disease-disease network (DDN) represents diseases as nodes and connections between diseases, such as observed or quantified biological factors, as edges^4^.

With the extensive growth of large-scale biomedical data, electronic health record (EHR)-linked biobanks have become a vital resource in the study of pleiotropy and the genetic architecture of complex traits. A phenome-wide association study (PheWAS) applied to an EHR-linked biobank can find hundreds of thousands of associations between phenotypes, such as diseases, clinical symptoms, or laboratory measurements, and genetic variants, such as common single-nucleotide polymorphisms (SNPs)^5^. Furthermore, PheWASs are disease- and variant-agnostic, meaning that the identification of these potential instances of pleiotropy remains unbiased^6,7^. The summary statistics from a PheWAS can be used to create corresponding shared-SNP DDNs (ssDDNs), where edges represent sets of associated SNPs that pass a desired threshold of significance and are shared between the two phenotypes^8–10^. By analyzing a ssDDN, a researcher or clinician can evaluate how diseases are linked to one another, with immediate insight into potential shared genetic architecture through the identification of putative pleiotropic SNPs at specific genomic locations.

EHR-linked biobanks often report quantitative lab results of blood- and urine-based biochemical markers. Many of these traits have a strong genetic basis, and they can be used as intermediate phenotypes in the analysis of complex diseases, offering additional information in the investigation of disease connections^11–15^. Given the polygenic predictive power of such continuous endophenotypes, integrating them into studies of non-mendelian disorders allows for improved interpretability at the molecular level, beyond what genetic pleiotropy can uncover^16^. Several individual laboratory measurements have been shown to be clinical predictors of cardiovascular disease, and evidence is accumulating for quantitative biomarker links with many other types of common diseases^17^. For example, Veturi et al. recently showed substantial pleiotropy between plasma lipids and diseases across many organ systems^18^. This is supported by over a decade of research from the Global Lipids Genetics Consortium, which has found that heritable lipid levels, such as lipoprotein cholesterols, triglycerides, and total cholesterol, are not only genetically related to complex diseases through shared loci, but are modifiable risk factors of those diseases^19–21^.

Based upon the additional insight that may be derived from such intermediate phenotypes, we propose a novel augmented version of the ssDDN, denoted as ssDDN+. Additional genetic associations between diseases are incorporated into the original ssDDN based upon shared genetic correlation with clinical laboratory measurements. We hypothesize that a ssDDN+ can represent inherited factors contributing to cross-phenotype associations and provide insight into the role of endophenotypes in these disease interactions. In this study, we constructed a ssDDN+ using PheWAS summary statistics from the UK Biobank (UKBB), revealing hundreds of genetic correlations between disease phenotypes and quantitative traits. We show that our augmented network uncovers genetic associations across different disease categories, connects relevant cardiometabolic diseases, and identifies specific biomarkers that are associated with the genetic architecture of multiple diseases. Comparing our ssDDN+ to its corresponding ssDDN demonstrates the complementary information that is revealed in this new network topology, highlighting the influence of quantitative traits within the diseasome^4^.

## RESULTS

### Disease networks

Disease-disease networks (as described in the Methods) were constructed using a cohort of approximately 400,000 participants from the UKBB for 318 binary disease phenotypes (Supplemental Table 1). The shared-SNP DDN, built using disease-SNP associations from PheWAS summary statistics alone, was augmented through the addition of edges representing shared genetic correlations with 31 clinical laboratory measurements (Fig. 1). These quantitative traits represent intermediate phenotypes which gain further insight into the genetics of binary disease traits (Supplemental Table 2). The unsigned, unweighted endophenotype-augmented disease network (ssDDN+) includes all edges from the baseline ssDDN, while incorporating additional genetic information from biomarkers.

**Fig. 1:**
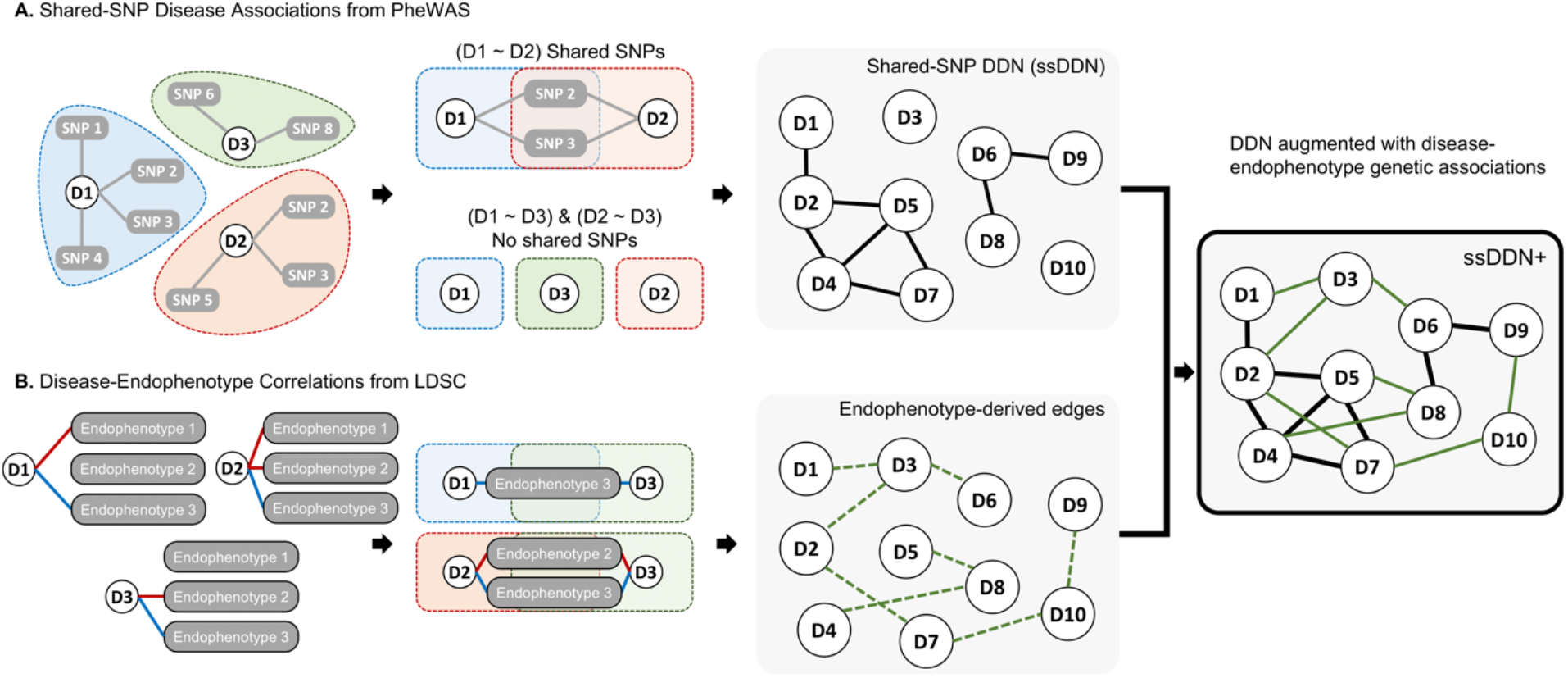
Overview of network construction. An overview of the process of developing the ssDDN+. (A) Disease phenotypes sharing genome-wide significant SNPs uncovered via a PheWAS are used to construct a shared-SNP DDN, where edges represent shared associations with variants between a pair of diseases. (B) Genetic correlation is determined between all diseases and quantitative endophenotypes, and if diseases are both genetically correlated (green dashed line) with the same endophenotype then edges between those diseases are added to the ssDDN.

### Additional edges in the ssDDN+

Using 322 significant genetic correlations between binary diseases and continuous measurements (Supplemental Fig. 1, Supplemental Fig. 2), we constructed a corresponding ssDDN+ from our UKBB ssDDN (Fig. 2). 1,561 new cross-phenotype genetic associations were identified compared with the original ssDDN, increasing the network’s total edge count by 242% (Supplemental Table 3). The ssDDN and ssDDN+ exhibited similar clustering behavior to one another (Supplemental Table 4). However, including indirect edges increased the connected node count from 114 to 138, meaning that 24 diseases gained connections to others because of associations derived from laboratory measurements. 116 indirect edges represented the same cross-phenotype associations as pre-existing direct edges, suggesting that highly significant SNPs associated with disease associations may be involved in the same pathways as the biomarkers that connect them. Indirect edges that contributed new information in the ssDDN+ can be explored online through our Human-Disease Phenotype Map browser at hdpm.biomedinfolab.com/ddn/biomarkerDDN. Additional network statistics for each DDN can be found in Supplemental Table 4.

**Fig. 2:**
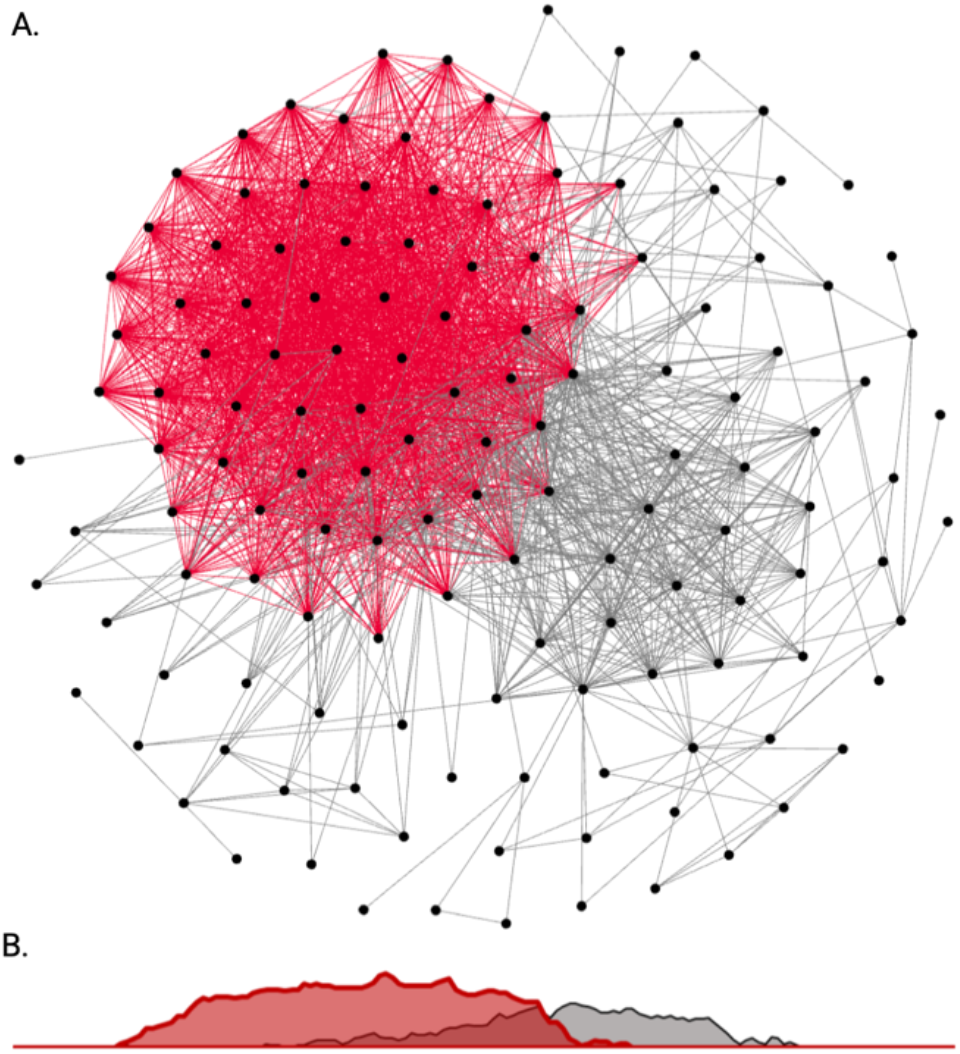
Endophenotype augmented disease-disease network. (A) A depiction of the full ssDDN+ based upon PheWAS summary statistics of binary disease phenotypes and continuous biomarker measurements from the UKBB. Gray edges represent direct shared-SNP edges, and red edges represent indirect biomarker genetic correlation edges. (B) A density plot projection of direct and indirect edge distributions in a single dimension. Direct and indirect edges identify different sets of genetic associations between diseases.

### Highly connected diseases and hub nodes

Within each DDN, a node’s degree, the number of other nodes to which it is connected, represents how genetically associated the corresponding disease is to other diseases. Hub nodes, nodes with the highest centrality in the graph, represent the most highly connected diseases. When we transition from the ssDDN to the ssDDN+, the relative degree of many diseases changes substantially. Fig. 3 demonstrates how the degree rank of disease phenotypes changes by supplementing the ssDDN with indirect edges and highlights known biology and genetic susceptibility for certain diseases. For instance, hyperlipidemia, a disease whose signal in our data is mostly represented by patients with hypercholesterolemia, has known mendelian effects from genes including *LDLR, APOB*, and *PCSK9*^22^. Correspondingly, we see hyperlipidemia has the top degree rank in the ssDDN. Furthermore, hyperlipidemia also exhibits known associations with a variety of lipidomic biomarkers^23^, justifying its role as the disease with the highest degree in the ssDDN+.

**Fig. 3:**
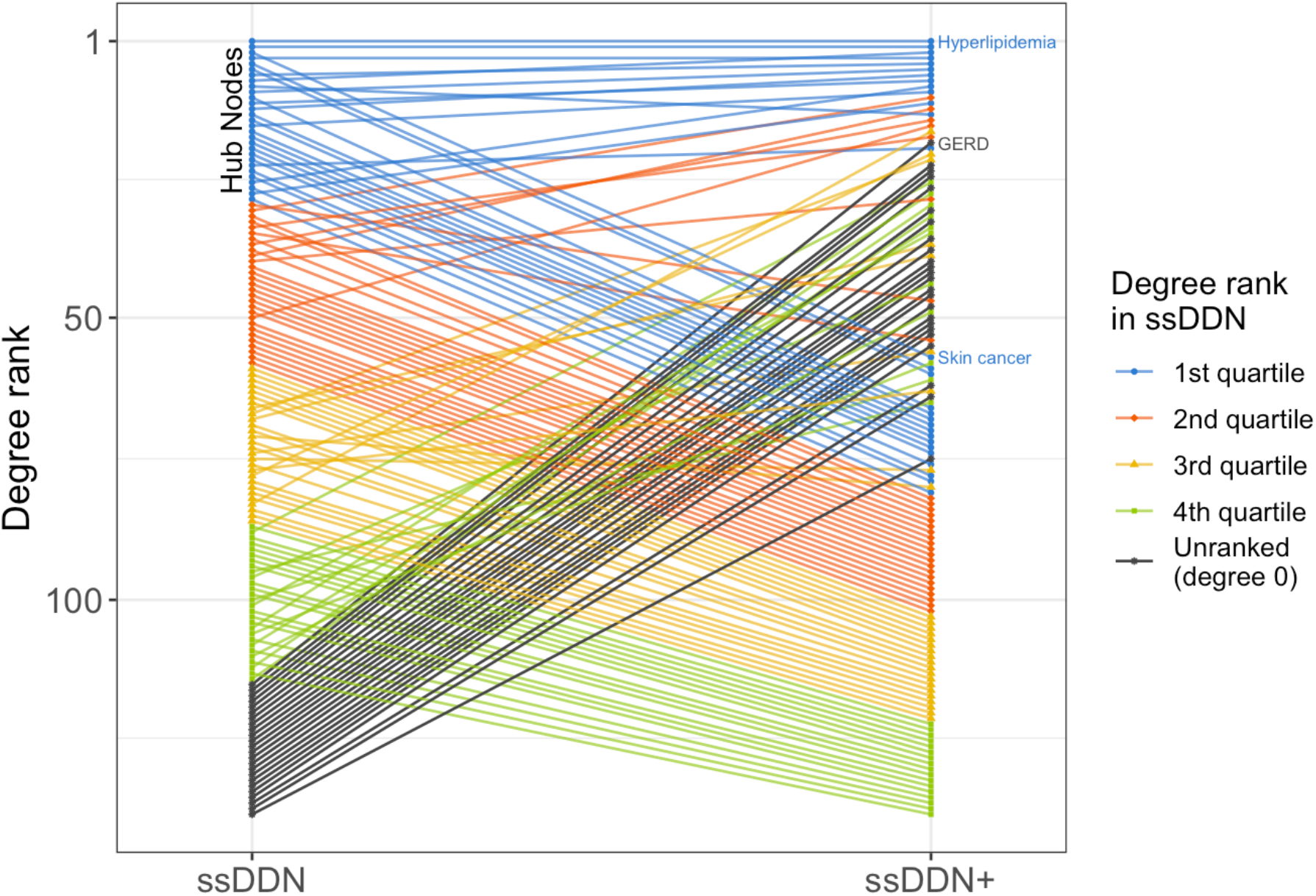
Change of node degree rank from ssDDN to ssDDN+. A slope graph of degree rankings for diseases in the ssDDN and ssDDN+. The degree of a node in a graph represents the number of other nodes to which it is connected. Within each network, degrees were computed for each node, and then diseases were ranked with respect to one another according to degree value. A rank of 1 represents the most connected disease. For both DDNs, hyperlipidemia (phecode 272.1) has the highest node rank. Ranks in the figure are colored by quartile within the ssDDN, with black representing nodes that became connected in the ssDDN+ after not having any connections in the original ssDDN. Some newly connected nodes (e.g., GERD) are hub nodes in the ssDDN+, while some highly connected nodes (e.g., skin cancer) became relatively less connected.

Many newly connected diseases also gain a high degree rank compared to other diseases after the inclusion of endophenotypes to the ssDDN. For instance, gastroesophageal reflux disease (GERD) has a known heritability estimate of roughly 31% based upon twin and family studies, with known risk genes including *FOXF1, MHC*, and *CCND1*^24^. However, the original ssDDN fails to capture any sort of genetic signal for GERD, meaning that the disease remains unconnected to other nodes. This failure to identify cross-phenotype associations with GERD in the original ssDDN is likely due to a combination of stringent significance thresholds for disease-variant association as well as external factors outside of genetics mitigating the associations that would otherwise be apparent in the input PheWAS data. Given known evidence that predictive biomarkers, such as C-peptide and TNF-alpha^25^, exist for GERD, this disease becomes a perfect candidate for the identification of additional information in the ssDDN+. Indeed, based upon the inclusion of endophenotypes, GERD gains one of the highest degree ranks in the ssDDN+.

Finally, some diseases that are originally hub nodes in the ssDDN become comparatively less influential in the ssDDN+. For instance, skin cancer is a hub node in the ssDDN and is a disease known to have common genetic associations with a variety of other neoplasms^26^. However, there is still a high amount of uncertainty in their clinical utility of biomarkers for skin cancer prognosis^27^. This behavior is reflected in both networks in our study. Within the ssDDN, skin cancer has a prominent position with respect to other diseases in the network. However, based upon the measures that we were able to incorporate into our ssDDN+, no additional edges are included for skin cancer. In other words, this specific ssDDN+ provides no further information about cross-phenotype associations with skin cancer as compared to its corresponding ssDDN.

### Differential contribution of endophenotypes by phenotype category

Although the addition of new edges in the ssDDN+ changes the topology of the network, this change is not evenly distributed across organ systems and disease types. Fig. 4a depicts specific pairs of phenotype groupings that become increasingly connected to one another by these new edges. In particular, a high concentration of new edges between the musculoskeletal and endocrine/metabolic disease categories is observed. This behavior is corroborated by prior research indicating associations between musculoskeletal degradation and the onset of metabolic disorders^28^. On the other hand, disease categories such as neoplasms and sense organs continue to remain relatively disconnected to other groupings, confirming conclusions drawn regarding cross-phenotype associations across disease categories in previous studies^4,8^.These differences across disease categories are due in part to the types of diseases that are genetically associated with the clinical measurements for which we had data to use. Indeed, we observe noticeable changes in the proportion of edges connected to diseases depending on category in the ssDDN+ (Fig. 4b). The most notable difference is the relative doubling of links connected to the phenotypes in the musculoskeletal system. Additionally, the proportion of edges that connect diseases from different groups increases from 75% to 85%, suggesting that endophenotypes may be useful in identifying additional genetic associations between diseases of different categories.

**Fig. 4:**
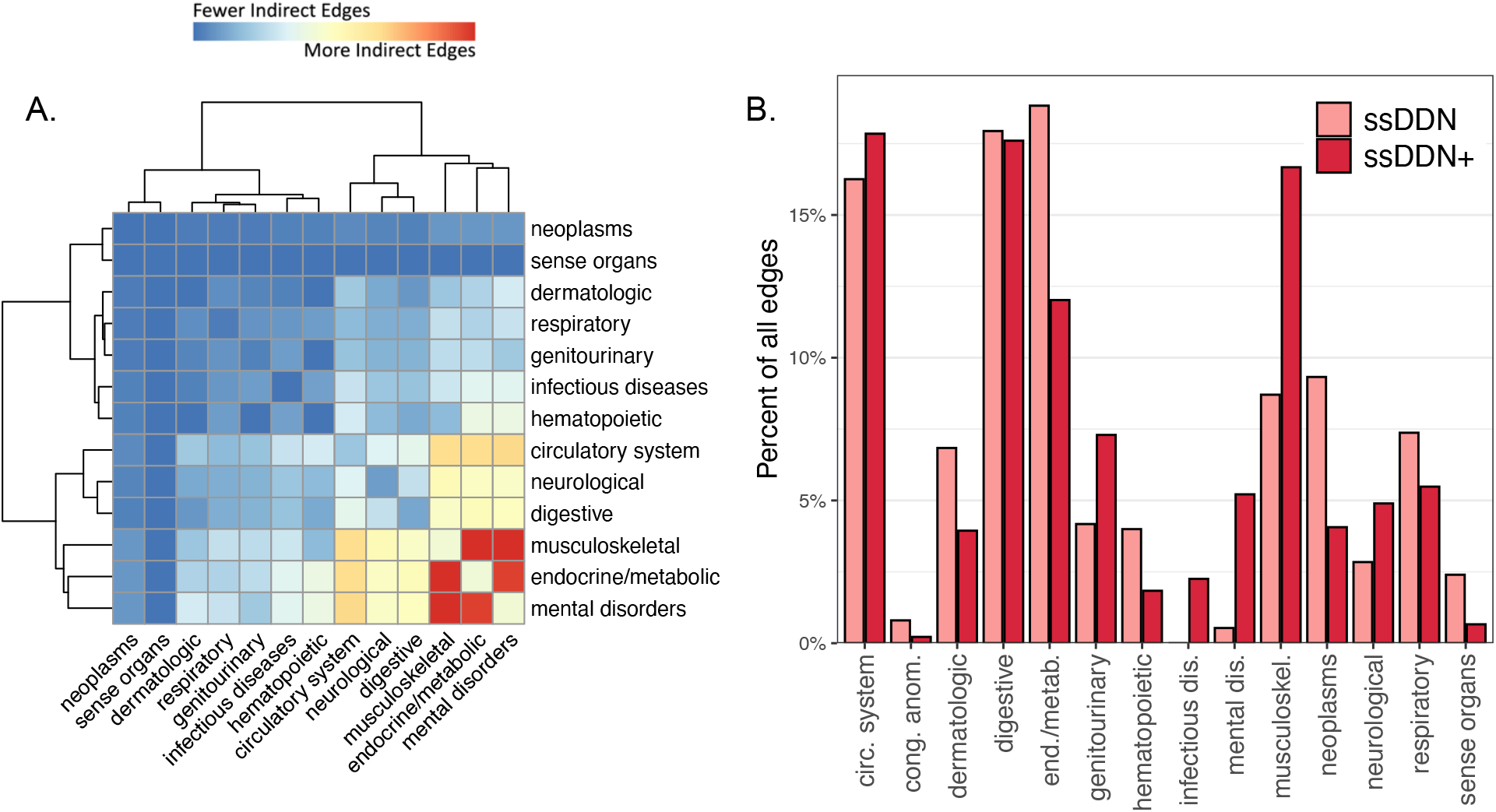
Category distribution of indirect edges. (A) A heatmap of the disease categories connected by indirect edges in the ssDDN+, normalized by the number of nodes in each category. Red represents category pairs with more indirect edges, and blue represents category pairs with fewer indirect edges. (B) A paired bar chart depicting the percentage of edges connecting at least one node in each disease category, colored by type of DDN. Some categories gain a disproportionately large number of edges from ssDDN to ssDDN+, while others gain only few edges.

### Cardiometabolic disease associations and influence of HDL-C

Previous research has highlighted a variety of potential genetic contributors to comorbidities among cardiometabolic diseases^29–31^, and an initial analysis of the ssDDN+ seems to confirm the influence of the endocrine/metabolic disease category. To further investigate such connections, we focus in on a subnetwork of our ssDDN+, where we consider only cardiometabolic phenotypes. The inclusion of 144 endophenotype genetic correlations increases the edge count from 116 to 200 when transitioning from the cardiometabolic ssDDN to its ssDDN+ (Supplemental Fig. 3). Multiple diseases of great interest, including heart failure, obesity, and type 1 diabetes, also become much more genetically connected, suggesting that in many instances, important disease connections may be missed in the ssDDN (Fig. 5).

**Fig. 5:**
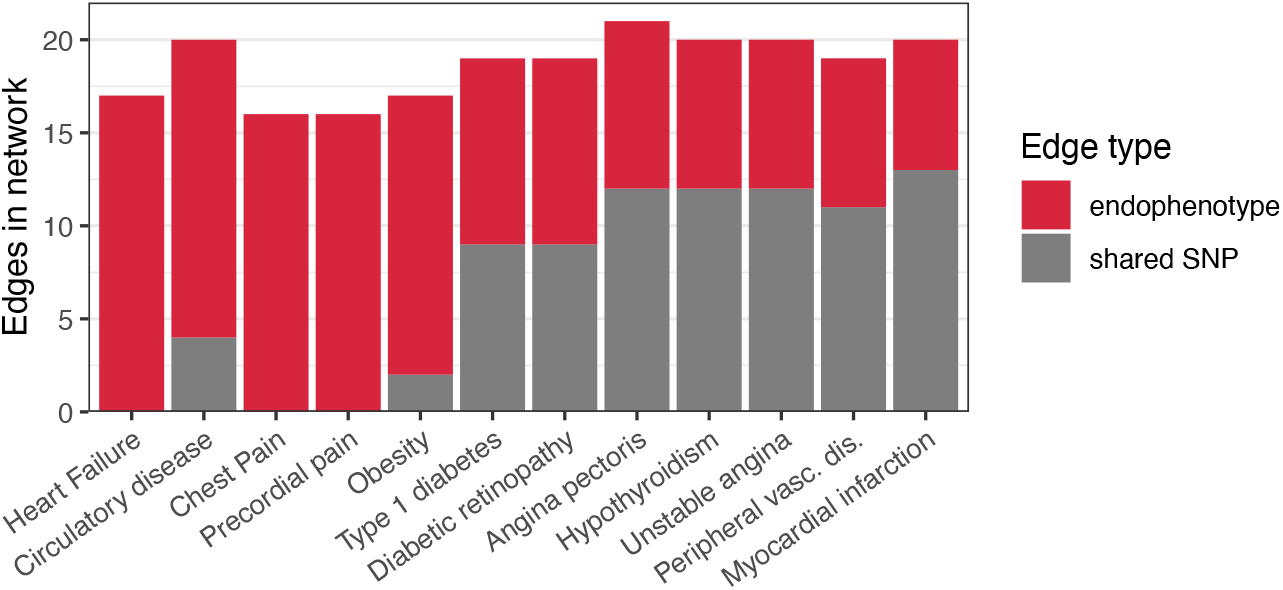
Cardiometabolic network edge types. A stacked bar chart depicting the types of links connected to the 12 disease phenotypes that gained the most edges going from the cardiometabolic ssDDN to the cardiometabolic ssDDN+. Gray represents direct shared-SNP edges, and red represents indirect endophenotype-correlated edges. Some diseases with known genetic drivers become connected to other phenotypes only as a result of indirect edges. For instance, clinical symptoms including heart failure, chest pain, and precordial pain, can only be connected to other chronic diseases after augmenting the ssDDN with endophenotypes.

The clinical traits used to build the ssDDN+ are involved in many different pathways, and thus we find certain biomarkers reveal many more edges than others. For instance, high-density lipoprotein cholesterol (HDL-C) contributes 996 new edges in the full ssDDN+ and 70 new edges in the cardiometabolic ssDDN+, while other biomarkers such as phosphates add no new edges (Supplemental Tables 5 and 6). This result highlights how clinical biomarkers may provide different levels of information from shared SNP links, and how phenotypes such as HDL-C may offer improved predictive power in identifying disease comorbidities. Focusing in on the cardiometabolic-specific ssDDN+, we can visualize how HDL-C adds considerable edges to the network. (Fig. 6). For instance, the inclusion of genetic correlation through HDL-C as edges connects hypothyroidism and angina pectoris, diseases known to be associated with HDL-C and with one another^32,33^.

**Fig. 6:**
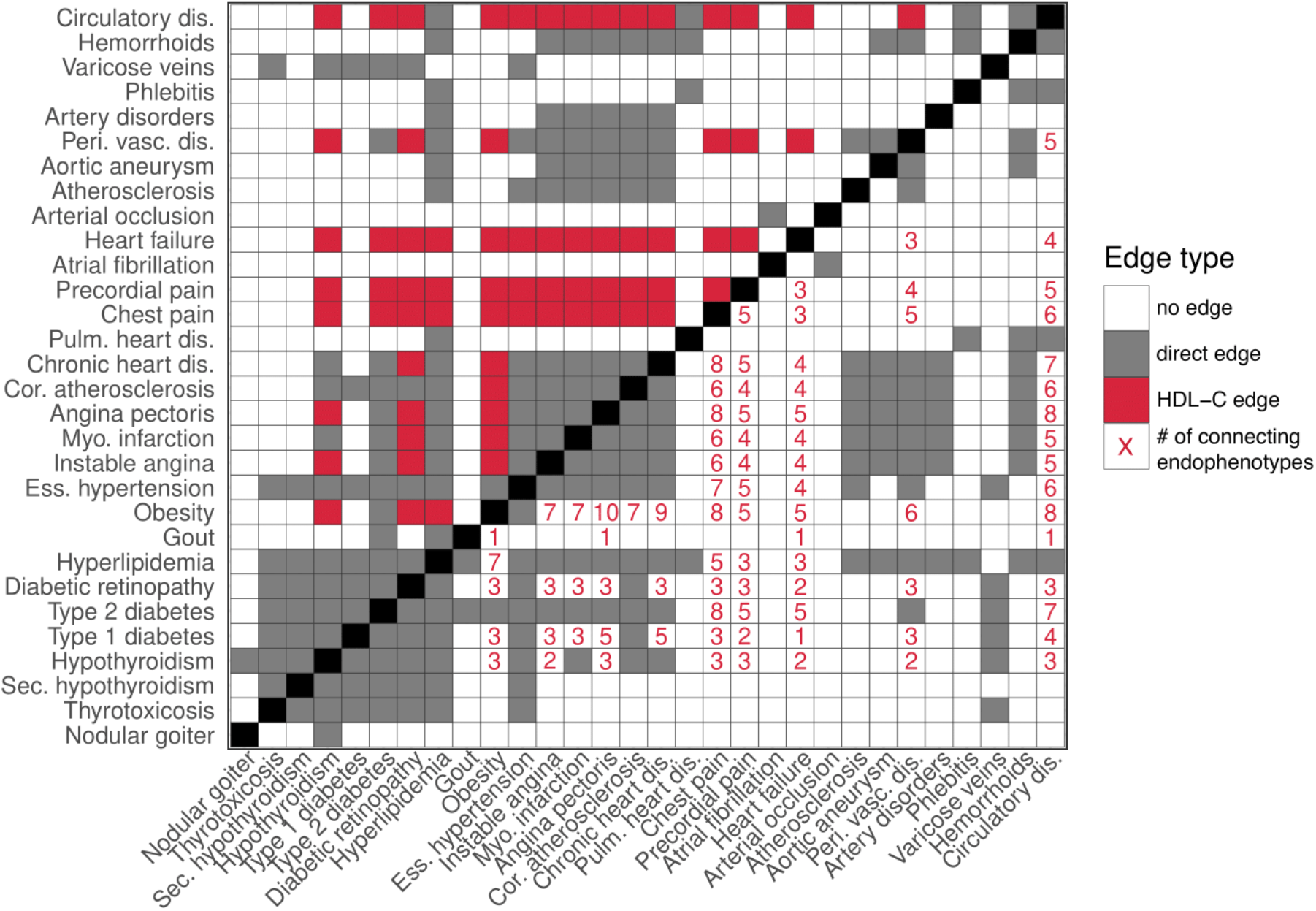
Contribution of edges by individual endophenotypes. An adjacency matrix presenting the contribution of HDL-C to edges in the cardiometabolic ssDDN+, as well as the influence of different clinical traits on connections among diseases. Gray squares represent associations identified between diseases through the shared SNP approach, while white squares represent a lack of connection between phenotypes. In the upper triangular adjacency matrix, red squares represent disease associations generated as a result of HDL-C. In the lower triangular matrix, red numbers represent the number of biomarker correlations shared between diseases in the ssDDN+.

Many of the endophenotype-disease associations found from our DDNs are validated by other genetic and epidemiological studies. Specifically, out of the 31 clinical measurements under consideration, HDL-C connects the greatest number of nodes. Furthermore, it has the strongest genetic correlation with type 2 diabetes (*r*_g_= 0.49). There is some evidence of HDL-C’s causal role in complex diseases, such as type 2 diabetes and coronary artery disease (CAD)^34–38^. Furthermore, the prominence of HDL-C and other fat-related compounds in our network, such as Apolipoprotein A and Triglycerides, is backed by a wealth of literature on their important roles in disease pathways. Additionally, HDL-C is known to have an association with increased risk of age-related macular degeneration^39^. Although macular degeneration was not included in our analysis, HDL exhibited a strong genetic correlation of 0.46 with a very similar phenotype that was considered – diabetic retinopathy. We also saw a strong association between triglycerides, another blood lipid that added a substantial number of edges to the ssDDN, and risk of CAD.

## DISCUSSION

In this study, we generated and analyzed a disease-disease network of genetic associations between binary phenotypes using significant SNPs from PheWAS summary statistics and genetic correlations with clinical laboratory measurements. Our network complements others by uncovering cross-phenotype links through genetic correlations between diseases and biomarkers, creating a denser model of the phenome. We highlighted disease classes as well as specific diseases with known genetic risk which benefit from this type of representation.

Studies of missing heritability throughout the past decade have made it apparent that considering only highly significant GWAS SNPs will often fail to capture the entire genetic architecture of complex diseases^40,41^. It is additionally important to functionally assess genetic effects – understanding the association between diseases and the disruption of molecular pathways through mutations can bring us closer to fully comprehending how diseases manifest as comorbidities and complications^42^. Both points highlight the utility of incorporating disease-associated biomarkers into the formation of human disease networks. Furthermore, PheWASs based on logistic regression binarize complex diseases that may have a range in their physical manifestation, making the use of endophenotypes even more pertinent.

In our analysis, the endophenotypes we incorporated contribute a non-random distribution of edges to specific diseases categories – musculoskeletal diseases gain more connections, while neoplasms gain much fewer. This difference is driven in part by which types of diseases have significant correlation with the biomarkers under consideration, and how for some phenotypes, the analysis of biological molecules is more useful when assessing genetic contributors. The differential augmentation across diseases provides evidence of the importance of including quantitative laboratory measurements. When looking specifically at cardiometabolic phenotypes, new edges are added from associations with biomarkers to phenotypes that have been determined to have strong polygenic causes, including heart failure^43^, obesity^44^, and diabetic retinopathy^45^.

The fundamental value this ssDDN+ adds is a novel way to model the diseasome. By harnessing the value of intermediate disease phenotypes, we can represent an increased number of genetic associations present in disease connections. For example, this ssDDN+ links Heart Failure (phecode 428.2), a disease with no connections in the ssDDN, to 54 other diseases. By integrating genetic correlations with endophenotypes into the ssDDN, we pick up additional signal that make the investigation of this and other phenotypes’ connections possible. Our network has multiple potential future applications, including drug design with network pharmacology, finding genetic targets for future therapeutics, and the advancement of personalized medicine and disease risk prediction^46^.

There are a few limitations to consider in our study. Though the binary diseases and continuous laboratory measurements both come from the UKBB, the summary statistics for each category of traits were generated by different groups with different processing conditions, yielding slightly different numbers of individuals in each case^47,48^. Within the binary disease PheWAS however, each GWAS uses a slightly different number of samples due to phenotype-specific exclusion criteria. Therefore, these relatively small differences in samples across PheWASs should not undermine our results. The two PheWASs also use slightly different criteria to define their SNPs, with one having around 13.7 million SNPs tested compared to roughly 28 million SNPs in the other. Since we harmonized the SNPs down to a count of 1.2 million variants with precomputed LD scores, this distinction does not impact our analysis. Additionally, we used very strict significance thresholds, both for finding shared SNPs between diseases and for determining legitimately genetically correlated biomarker-phenotype pairs. Although this stringency may result in missing some genetic associations between diseases, it allows us to be confident in the connections we do observe in the ssDDN. We also note that our DDNs represent data only for the UKBB population, meaning that conclusions drawn from our analysis can only be interpreted from a British European perspective. Finally, despite the fact that the phecode system of disease classification is more aligned with definitions of biomedical research than comparable disease encoding systems such as ICD-9 or ICD-10^49^, we appreciate that it is still imperfect at capturing true occurrences of phenotypes in patients. As a result, any conclusions drawn from our analysis need to bear this potential inaccuracy in mind.

In conclusion, we built an augmented disease-disease network that integrates genetic correlations with endophenotype measurements to represent additional cross-phenotype associations. Further steps in our analysis involve considering additional clinical traits depending on data availability, as well as additional population cohorts, as we may find even more endophenotype associations and thus more network edges^50^. We also hope to compare ssDDN+s to their corresponding ssDDNs given different significance thresholds for associations between diseases and SNPs. Future work should consider integrating mendelian randomization to identify the causality behind the correlative relationships that were uncovered. Additionally, full analysis of networks built from all levels of diseases risk (SNP-based, gene-based, symptom-based, molecular-based, pathway-based, microRNA-based, exposure-based, etc.) will be essential to integrate into studies and patient-prediction tasks^51^, along with multilayer graphs that summarize complex biological architecture beyond individual edges. Overall, our method helps to navigate the study of complex diseases and enables further network-based analysis involving pleiotropy, polygenicity, and heterogeneity. Our results can facilitate future network-based research of diseases, uncovering potential sources of missing heritability in multimorbidities and highlighting potential genetic targets for precision medicine investigations.

## METHODS

### Phenome-wide association analyses

PheWAS summary data from the UKBB were used to investigate the genetic relations among disease phenotypes. To derive genetic associations for binary disease phenotypes, a PheWAS was run for 400,000 British individuals of European ancestry with 1,403 phecode-labeled phenotypes using SAIGE^52^, controlling for sex, age, genetic relatedness, and the first four principal components^48^. Imputation using the Haplotype Reference Consortium panel yielded 28 million imputed SNPs, with all genomic positions on GRCh37^53^. To improve interpretability of diseases under consideration, we removed phenotypes if they had a case count less than 1,000 cases, had a phecode encoding specific to the hundredths digit, or belonged to phecode categories of “symptoms” or “injuries & poisonings”. Additional manual curation was also applied to remove hierarchically related diseases with similar case counts that would have represented correlated phenotype signals, resulting in a final set of 318 binary phenotypes. To derive genetic associations for continuous endophenotypes, a PheWAS was run for 361,194 British individuals of European ancestry with 31 rank-normalized quantitative biomarker measurements (Supplemental Table 2). This PheWAS was performed for 13.7 million QC-passing SNPs using Hail 0.2^54^, corrected for sex, age, and the first 20 principal components^47^. Between the two PheWASs, for alleles to remain consistent across the full set of diseases and biomarkers, variants were restricted to a unified list of HapMap3 SNPs. Due to the complicated LD structure in the major histocompatibility complex, SNPs in that region were also removed^12,55^. As a result, roughly 1.2 million SNPs remained for the identification of associations between diseases and laboratory measurements^56^.

### Disease-endophenotype correlations

The shared-SNP approach of identifying genetic associations between traits is a reasonable assumption for binary traits given the shared components hypothesis^2^. However, in the case of evaluating genetic associations between binary traits and continuous traits, such a method may fail to appropriately capture the magnitude of associations with the quantitative marker. Linkage disequilibrium score regression (LDSC)^57^ offers an effective method of calculating genetic correlations between pairs of phenotypes through the analysis of PheWAS summary-level data^58^. This process considers all common SNPs in a genome regardless of significance, accounting for SNP weight when determining associations between traits.^12,59^. Applying LDSC to the summary statistics described in the above section, we generated bivariate genetic correlation values (*r*_*g*_) between each binary disease phenotype and each quantitative endophenotype. Filtration to consider only genetic correlations for heritable phenotypes produced 9,566 disease-endophenotype *r*_*g*_ estimates. Of these correlations, 322 were found to be significant with a false discovery rate (FDR) < 0.05^12,60,61^.

### Construction of ssDDN and ssDDN+

Curated PheWAS summary data for 318 binary traits were used to generate the baseline ssDDN. The augmented version of the ssDDN, the ssDDN+, was constructed by incorporating the same PheWAS summary data with genetic correlations between the 318 binary traits and 31 additional continuous traits (Fig. 1). The methodology described by Verma et al.^8^ was applied to create the ssDDN. An edge in the set **E** = {e_ij_}^|***V***|×|***V***|^ was established between each pair of binary phenotypes v_i_ and v_j_ if the two diseases shared associations with at least one common SNP at a genome-wide significance threshold of 5 × 10^−8^.^9,10,62,63^ e_ij_ represents the presence or absence of a connection, meaning that e_ij_ = 1 if v_i_ and v_j_ had any common shared SNPs and e_ij_ = 0 otherwise. These edges can be thought of as direct links between phenotypes in the ssDDN. The final ssDDN is an undirected, unweighted graph.

The corresponding ssDDN+ can be represented as graph 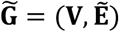, where node set **V** represents the set of binary phenotypes and edge set 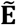 represents all connections between phenotypes. 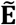 can be decomposed into direct connections (**E**) obtained from the ssDDN and indirect connections (**E**^+^)estimated from significant genetic correlations derived from LDSC. We constructed a genetic correlation matrix **R =** {r_ij_ ∈ ℝ}^|***V***|×|***T***|^ where ***T*** represents the set of all quantitative traits. The correlation matrix **R** was transformed into an association matrix 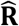, such that 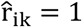 if the genetic correlation r_ik_ between phenotype v_i_ and quantitative trait t_k_ passed statistical significance. Then, the indirect connection 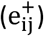 was established by determining whether phenotypes v_i_ and v_j_ shared a genetic association with the same trait t_k_ with 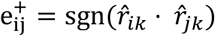, where sgn(·) is the signum function. If a common genetically correlated quantitative trait was identified between the two phenotypes, then an indirect edge was included in **E**^+^. The final graph, with edge set 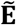 comprising of the union of **E** and **E**^+^, corresponds to the complete undirected, unweighted ssDDN+. To generate, visualize, and analyze both graphs, we made use of Gephi 0.90^64^ and sigma.js^65^, open-source network visualization software packages, as well as NETMAGE^10^, a web-based tool that allows users to upload PheWAS summary statistics and generate corresponding interactive disease-disease networks. Further analysis and visualization of DDN network statistics were performed using R 4.1.3^66^.

### Disease categories

The 318 phecode-encoded binary phenotypes were organized into 15 unequally sized categories (Supplemental Table 1)^67^. Category-specific analyses allowed us to assess how the network structure of the ssDDN+ can provide insight into connections between biologically similar diseases that affect the same organ systems. We considered phenotypes in either the groups “endocrine/metabolic” and “circulatory system” as cardiometabolic diseases.

## Data Availability

All data produced are available online at https://www.leelabsg.org/resources.

## DATA AVAILABILITY

Summary statistics for binary disease phenotypes can be accessed at https://www.leelabsg.org/resources and for continuous laboratory measurements at https://nealelab.github.io/UKBB_ldsc/downloads.html. Additionally, new endophenotype disease connections can be explored on our web visualization tool at https://hdpm.biomedinfolab.com/ddn/biomarkerDDN.

## ACKNOWLEDGEMENTS

This work has been supported by the National Institute of General Medical Sciences (NIGMS) R01 GM138597 and S10OD023495.

## Author contributions

JW, VS, YN, and DK were involved in designing and conceptualizing the study. DK supervised the study. JW and VS performed data curation and analysis. JW and VS wrote the original manuscript. All authors revised and approved the final manuscript.

