## Supplemental information for "Uncovering genetic associations in the human diseasome using an endophenotype-augmented disease network"

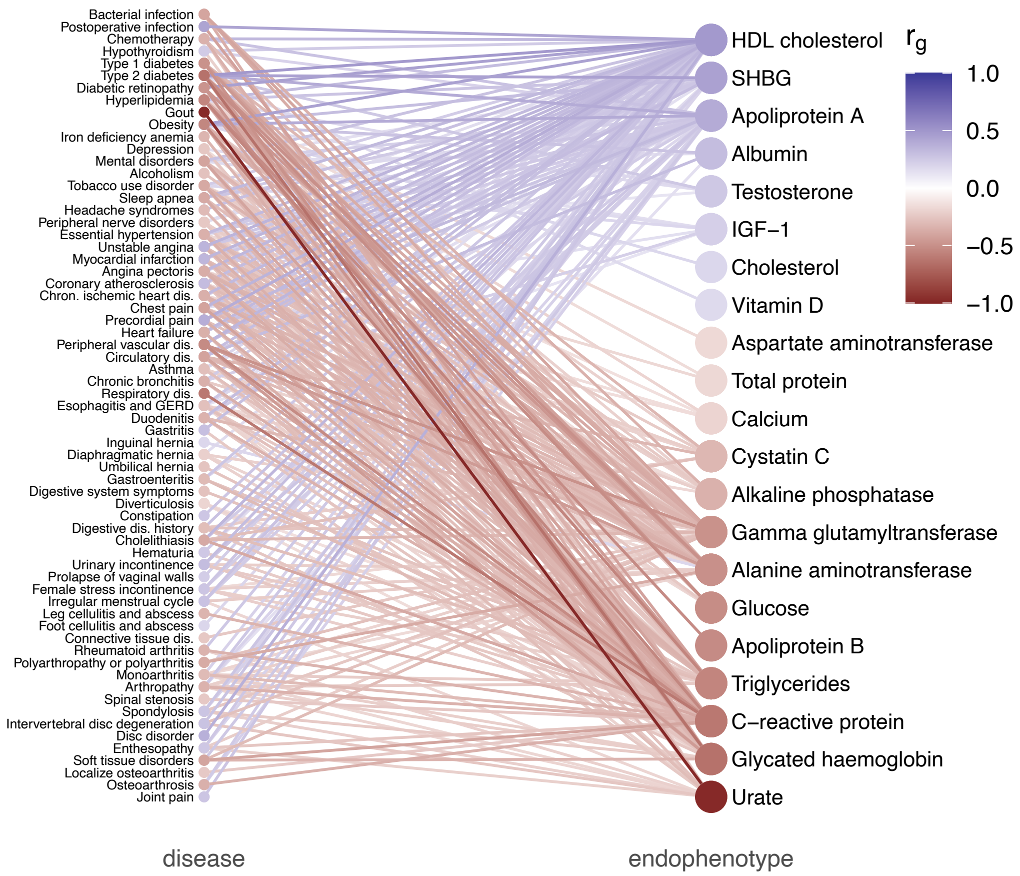


**Supplemental Fig. 1: Bipartite graph of genetic correlations between diseases and endophenotypes**

A bipartite graph showing all 322 genetic correlations between binary disease phenotypes and quantitative measurements. Edges are colored according to correlation value.

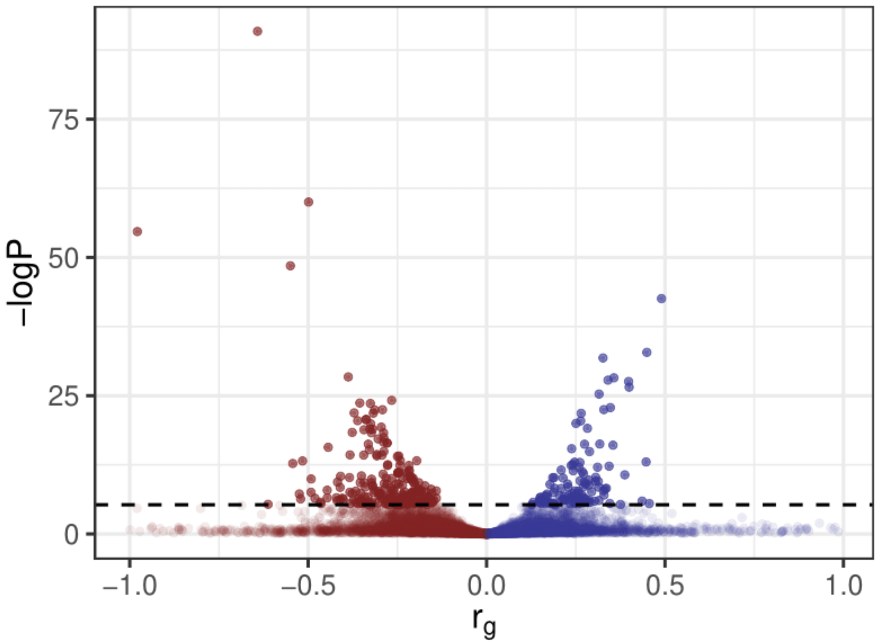


**Supplemental Fig. 2: Distribution of genetic correlations between binary diseases and endophenotypes**

A volcano plot showing significance level against $r_g$ for all computed genetic correlations between heritable clinical measurements and disease phenotypes. The 322 significant genetic correlations are highlighted, along with the threshold of the correction factor (FDR < 0.05). There is an even distribution of positive and negative correlations.

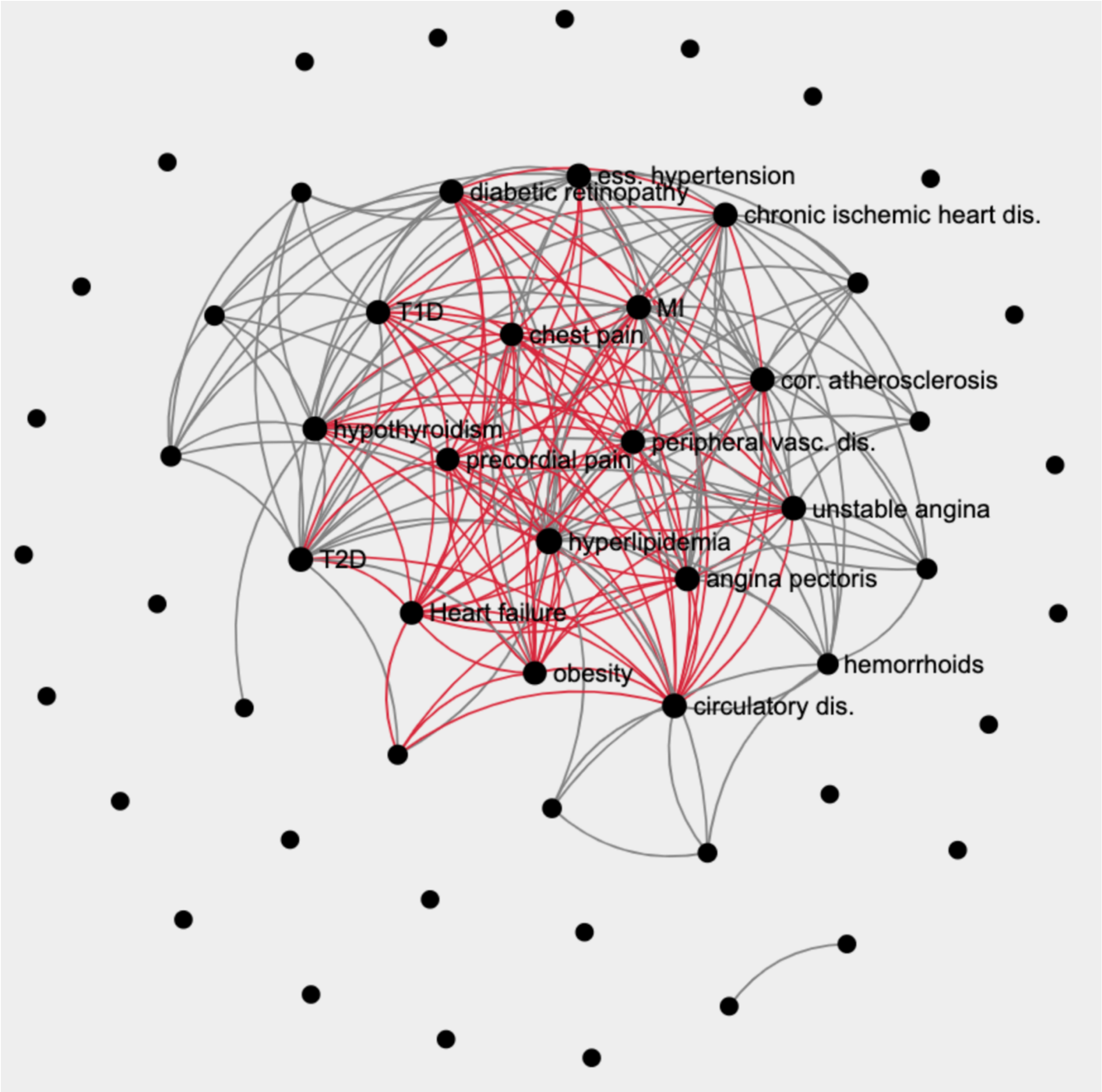


**Supplemental Fig. 3: Cardiometabolic ssDDN+**

A subnetwork of the full ssDDN+ including only endocrine/metabolic and circulatory system diseases, colored by edge type: gray for direct and red for indirect. Phenotypes gaining the most edges through genetic correlation with quantitative endophenotypes are labeled in the network.
